# Financial factors: a mixed methods survey-based study of barriers and facilitators to physical activity in type 1 diabetes

**DOI:** 10.64898/2025.12.18.25342586

**Authors:** Kara C. Anderson, Sara Ann Mauro, Alayna A. Panzer, Daria Igudesman, Kara S. Fitzgibbon, Shayne Zaslow, Kaitlin M. Love

## Abstract

**Aims:** To identify barriers and facilitators to physical activity (PA) in adults with type 1 diabetes (T1D) living in the United States (U.S.) and identify sociodemographic factors related to meeting recommended PA.

**Methods:** We conducted a cross-sectional online survey study of adults with T1D aged ≥18 years recruited through online-based platforms. Quantitative questions related to exercise quantity and intensity, demographic characteristics, and exercise barriers and facilitators. Wilcoxon rank sum tests or independent t-tests were used to compare quantitative responses in individuals meeting or below target PA. Barriers and facilitators were also assessed qualitatively with open-ended questions. Logistic regression was performed to determine if the following characteristics were independently associated with meeting PA recommendations: age, sex, income level, and automated insulin delivery system use.

**Results:** Of 281 respondents who completed questions about exercise quantity, 162 (57.7%) were women, mean age 52.6 ± 16.6 years, and 151 (53.7%) met PA guideline recommendations. Common barrier themes related to T1D included hypoglycemia, time, lack of knowledge about glycemic management, cost, and failure of available treatments to accommodate exercise. Common facilitator themes were insurance reimbursement of exercise program/facility, peer exercise groups, health/fitness advising, and T1D tailored fitness. Middle (vs. upper) income level was independently associated with lower odds of meeting PA recommendations (adjusted odds ratio 0.46, 95% CI: 0.27, 0.78, p = 0.004).

**Conclusions:** In this predominately U.S. cohort with T1D, financial factors were common novel themes related to PA. Further validation in more socioeconomically diverse cohorts and research examining PA reimbursement cost-efficacy are needed.

**Novelty statement:** *What is already known?:*

- In prior qualitative studies in type 1 diabetes, hypoglycemia is a commonly reported barrier to physical activity (PA) engagement. Most studies were conducted outside the United States (U.S.).

*What this study found:*

- In a predominately U.S. cohort of adults with type 1 diabetes, cost is a newly identified barrier to PA.
- Insurance reimbursement of PA programs/facilities was a reported facilitator.
- Individuals with highest income were 54% more likely to achieve recommended PA compared to other income categories.

*What are the implications of the study?:*

- Cost-efficacy research examining PA programs/facility reimbursement in type 1 diabetes is needed.

## Introduction

Type 1 diabetes increases cardiovascular disease mortality risk despite individuals maintaining target glycemic control. Cardiovascular disease is the leading cause of death following 20 years of type 1 diabetes, and type 1 diabetes shortens life span by 9 to 17 years (1,2). Earlier type 1 diabetes onset exaggerates risk of cardiovascular disease in this population and worsens years of life lost (2). In type 1 diabetes, interventions to improve longevity through cardiovascular disease prevention are needed.

For individuals with type 1 diabetes, physical activity confers important longevity benefits according to prospective longitudinal cohort studies. In the EURODIAB study, moderate to vigorous intensity physical activity at least once weekly was associated with a 34% lower risk of death, and it reduced the occurrence of cardiovascular disease events among women (3). In the Pittsburgh Insulin-dependent Diabetes Mellitus Morbidity and Mortality study, sedentary males with type 1 diabetes were three times more likely to die prematurely than active males with type 1 diabetes, and a similar, though not statistically significant, association was found in females (4). The relationship persisted even after adjusting for traditional cardiovascular disease risk factors (4). These results were reinforced by the Finnish Diabetic Nephropathy (FinnDiane) study, a larger nationwide cohort study, in which individuals with type 1 diabetes also had a lower risk of all-cause mortality when participating in moderate to high levels of physical activity compared to low levels (5). Collectively, these observational studies strongly indicate a critical role for exercise in longevity—likely predominately mediated by cardiovascular disease prevention—in people with type 1 diabetes.

Short term mechanistic interventional studies further corroborate the observational data supporting cardiovascular benefits of exercise. Nitric oxide (NO)-dependent endothelial dysfunction, measured by flow mediated dilation, is an early marker of vascular dysfunction and a predictor of cardiovascular events; therefore, it is a key surrogate outcome measure for cardiovascular disease. In an intervention study implementing 16 weeks of moderate continuous exercise training, individuals with type 1 diabetes had improved flow mediated dilation (6).

Similarly, 8-18 weeks of high intensity interval training improved flow mediated dilation in other type 1 diabetes studies (7,8). It is important to note that these vascular benefits were not maintained after discontinuation of exercise (6). This underscores the importance of not only engaging in but also maintaining an exercise program. People with type 1 diabetes often experience other benefits from exercise which could provide indirect cardiovascular disease protection like improved mean glucose and time in target range (9). Despite their elevated cardiovascular disease risk and advantages of physical activity, individuals with type 1 diabetes exercise less on average than their unaffected peers (10,11). Reasons for this dissonance remain incompletely defined.

Prior qualitative research examining barriers and facilitators to physical activity in type 1 diabetes were predominately conducted in the United Kingdom, Canada, or Australia (12–16). There is currently limited literature focusing on the United States (U.S.) population (17), in which sedentary behaviors are increasing in the general population and where persons with type 1 diabetes are disproportionately impacted by high healthcare costs relative to those with type 2 diabetes or without diabetes (18,19). In a prior U.S. qualitative study of adults with type 1 diabetes, interview participants were physically inactive (17). Qualitative data are needed to bolster this limited evidence given that diabetes technologies such as automated insulin delivery (AID) systems and continuous glucose monitors (CGMs) that provide safeguards to protect against hypoglycemia surrounding exercise are costly in the U.S. This may lead to unique barriers in U.S. populations compared to countries with national healthcare coverage and/or subsidized CGM devices.

No published type 1 diabetes studies have simultaneously collected both qualitative and quantitative data regarding exercise quantity and intensity, diabetes quality of life, socioeconomic status and insurance status, and exercise facilitators and barriers. Most studies identified hypoglycemia or fear of hypoglycemia as a frequent barrier to physical activity in type 1 diabetes (12–14,17). Studies in type 1 diabetes have also not yet compared individuals who are meeting physical activity targets to those not meeting targets, which could be useful to identify key factors unique to this population which prevent or support physical activity implementation. This study seeks to address gaps in the literature regarding barriers and facilitators to physical activity among adults with type 1 diabetes recruited from U.S.-based online platforms through a comprehensive survey-based approach. Understanding the perspectives of adults with type 1 diabetes living in the U.S. will better identify and support interventions to improve physical activity engagement.

## Methods

### Oversight

This study was approved by the University of Virginia Social Behavioral Science Institutional Review Board (#7009).

### Eligibility/Units of Study

Adults, ages 18 years and older, with self-reported type 1 diabetes were recruited from Reddit via social media advertisement and the T1D Exchange via social media platform advertisement and Research Registry email. The T1D Exchange reaches an Online Community of roughly 35,000 subscribers, roughly 45,000 followers on its organizational social media platforms and has over 21,000 participants living with or caring for someone with type 1 diabetes in its online Registry.

### Data Collection Instrument and Methods

The survey instrument (Online Supplemental Material) was developed by building on validated quantitative and qualitative surveys in this population (12,13,20), refined with input from experienced researchers at the Center for Survey Research at the University of Virginia (K.F. and A.P.), and tested in clinical trial (NCT05478707) participants with type 1 diabetes to ensure culturally appropriate language adaptation. The survey was conducted online on the REDCap platform (2/10/2025-3/29/2025) at which point qualitative responses were assessed. Participants did not receive incentives.

### Quantitative Methods and Analysis

Quantitative survey questions elicited information about PA duration and intensity. Our PA categories were based on American Diabetes Association 2025 guidelines, which recommend 150 minutes of moderate intensity activity per week or 75 minutes/week of high intensity activity (21). Participants reported engaging in multiple intensities and duration of physical activity (PA), and PA was subsequently classified as either target PA or below target PA. Target PA was defined as performing any of the following PA in a typical week: 1) moderate intensity exercise ≥ 150 minutes/week, 2) high intensity exercise ≥ 70 minutes/week, 3) moderate intensity exercise ≥ 120 minutes/week and high intensity exercise ≥ 30 minutes/week, 4) moderate intensity exercise ≥ 90 minutes/week and high intensity exercise ≥ 30 minutes/week, or 5) moderate intensity exercise ≥ 9 minutes/week and high intensity exercise 50 – 69 minutes/week. Engagement in less PA than those categories was defined as “below target” PA. Information collected about demographic characteristics included age, sex, race, ethnicity, education level, insurance payor, and income level. Income level was defined by the following annual household income classifications: upper ≥ $150,000, middle $50,000 - $149,999, lower < $50,000. Diabetes-specific questions included duration of diabetes, diabetes technology use (i.e. use of any of the following: continuous glucose monitor [CGM], insulin pump without sensor augmentation, sensor augmented pump, hybrid closed loop system), and diabetes satisfaction using questions from the 6-item Diabetes Quality of Life Brief Clinical Inventory (20). Respondents rated exercise barriers using a 7-point Likert scale, ranging from extremely unlikely (1) to extremely likely (7) to prevent PA in the next three weeks.

### Analysis

Using R version 4.1.3, summary statistics were performed to examine demographic characteristics and responses to exercise facilitators and barriers. Wilcoxon rank sum tests or independent t-tests were used to compare quantitative responses in individuals meeting or not meeting target PA as appropriate. Multiple, multivariable logistic regression analysis models were performed to determine if the following covariates were independently associated with meeting guidelines-based PA: sex, age, education level, income level, automated insulin delivery (AID) system—defined as sensor augmented pump or hybrid closed loop system--or continuous glucose monitor (CGM) use.

### Qualitative Methods and Analysis

The questionnaire included five open-ended questions focused on the facilitators and barriers to exercise and how living with type 1 diabetes impacts exercise participation. The open-ended responses were exported to Microsoft Excel for qualitative content analysis (22). Coding and thematic analysis were performed by experienced qualitative researchers (A.P. and S.Z). For each question, all responses were reviewed at least twice, and codes were generated for frequently mentioned categories. After the initial coding, the team reviewed the coding to combine and separate codes, ensuring they accurately reflected the content of the responses.

## Results

Two hundred and eighty-one adults participated in this study and had complete PA responses (FLOW diagram in Online Supplemental Fig. 1) with mean age: 52.6 ± 16.6 years, %female: 57.7, and disease duration 31.2 ± 19 years. Within this sample, 53% were meeting the PA targets (n=151, age: 51.8 ± 17.1 years, %female: 58.3%, disease duration 28.5 ± 18.8 years). Sex, age, and diabetes duration were similar between those meeting and not meeting PA targets (below target: n=130, age: 53.7 ± 16.1 years, %female: 56.9%, disease duration: 34.4 ± 18.8 years). Full demographics are listed in **Table 1**. Of the respondents, 99.6% reported exercise is beneficial for people with type 1 diabetes.

**Table 1:** Demographic Characteristics by Not Meeting or Meeting Target Physical Activity.

|  | **Full Cohort**<br>**(n=281)** | **Not Meeting Target**<br>**Physical Activity**<br>**(n=130)** | **Target Physical**<br>**Activity**<br>**(n=151)** |
| --- | --- | --- | --- |
| Age (years) | 52.6 ± 16.6 | 53.7 ± 16.1 | 51.8 ± 17.1 |
| Female sex, no.<br>(%) | 162 (57.7%) | 74 (56.9%) | 88 (58.3%) |
| Diabetes duration<br>(years) | 31.2 ± 19 | 34.4 ± 18.8 | 28.5 ± 18.8 |
| Established with<br>endocrinologist—<br>no. (%) | 273 (98.6%) | 108 (98.2%) | 165 (98.8%) |
| CGM Use—no.<br>(%) | 229 (81.5%) | 106 (81.5%) | 123 (81.5%) |
| Diabetes<br>treatment—no.<br>(%) | MDI: 61 (21.7%)<br>Non-SAP: 16 (5.7%)<br>SAP: 56 (19.9%)<br>HCL: 142 (50.5%) | MDI: 24 (18.5%)<br>Non-SAP: 7 (5.4%)<br>SAP: 29 (22.3%)<br>HCL: 67 (51.5%) | MDI: 37 (24.5%)<br>Non-SAP: 9 (6.0%)<br>SAP: 27 (17.9%)<br>HCL: 75 (49.7%) |
| Race—no. (%) | American Indian/<br>Native Alaskan: 3<br>(1.1%)<br>Asian/Asian<br>American: 9 (3.2%)<br>Black/African<br>American: 5 (1.8%)<br>Mixed: 1 (0.4%)<br>White: 237 (84.3%)<br>Not reported: 26<br>(9.2%) | American Indian/<br>Native Alaskan: 0 (0%)<br>Asian/Asian American:<br>3 (2.3%)<br>Black/African<br>American: 4 (3.1%)<br>Mixed: 0 (0%)<br>White: 111 (85.4%)<br>Not reported: 12<br>(10.8%) | American Indian/<br>Native Alaskan: 3<br>(2.0%)<br>Asian/Asian American:<br>6 (4.0%)<br>Black/African<br>American: 1 (0.6%)<br>Mixed: 1 (0.6%)<br>White: 126 (83.4%)<br>Not reported: 14<br>(9.3%) |
| Hispanic/LatinX<br>ethnicity—no.<br>(%) | 5 (1.8%) | 1 (0.8%) | 4 (2.6%) |
| Location—no.<br>(%) | Rural: 43 (15.3%)<br>Suburban: 147<br>(52.3%)<br>Urban: 69 (24.6%)<br>Not reported: 22<br>(7.8%) | Rural: 18 (13.8%)<br>Suburban: 67 (51.5%)<br>Urban: 34 (26.2%)<br>Not reported: 11 (8.5%) | Rural: 25 (16.6%)<br>Suburban: 80 (53.0%)<br>Urban: 35 (23.2%)<br>Not reported: 11 (7.3%) |
| Education level—<br>no. (%) | High School/GED:<br>18 (6.4%)<br>Associate's degree:<br>43 (15.3%)<br>Bachelor's/Some<br>Graduate Level: 89<br>(31.7%) | High School/GED: 8<br>(6.1%)<br>Associate's degree:<br>20 (15.4%)<br>Bachelor's/Some<br>Graduate Level: 43<br>(33.1%) | High School/GED: 10<br>(6.6%)<br>Associate's degree:<br>23 (15.2%)<br>Bachelor's/Some<br>Graduate Level: 46<br>(30.5%) |
|  | Graduate degree: 110 (39.1%)<br>Unknown: 21 (7.5%) | Graduate degree: 48 (36.9%)<br>Unknown: 11 (8.5%) | Graduate degree: 62 (41.1%)<br>Unknown: 10 (6.6%) |
| Household income—no. (%) | Less than \$50,000: 25 (8.9%)<br>\$50,000- 149,999: 114 (40.6%)<br>Greater than or equal to \$150,000: 123 (43.8%)<br>Unknown: 19 (6.8%) | Less than \$50,000: 11 (9.9%)<br>\$50,000- 149,999: 51 (45.9%)<br>Greater than or equal to \$150,000: 39 (35.1%)<br>Unknown: 10 (9%) | Less than \$50,000: 12 (7.9%)<br>\$50,000- 149,999: 52 (34.4%)<br>Greater than or equal to \$150,000: 78 (51.7%)<br>Unknown: 9 (6.0%) |
| Insurance payor—no. (%) | Medicaid: 3 (1.1%)<br>Medicare: 73 (26%)<br>Non-US Universal: 12 (4.3%)<br>Private: 159 (56.6%)<br>Tricare: 3 (1.1%)<br>Uninsured: 4 (1.4%)<br>Unknown: 27 (9.6%) | Medicaid: 1 (0.8%)<br>Medicare: 36 (27.7%)<br>Non-US Universal: 3 (2.3%)<br>Private: 73 (56.2%)<br>Tricare: 2 (1.5%)<br>Uninsured: 2 (1.5%)<br>Unknown: 13 (10%) | Medicaid: 2 (1.3%)<br>Medicare: 37 (24.5%)<br>Non-US Universal: 9 (6.0%)<br>Private: 86 (57%)<br>Tricare: 1 (0.7%)<br>Uninsured: 2 (1.3%)<br>Unknown: 14 (9.3%) |

Abbreviations: CGM, continuous glucose monitor; HCL, hybrid close loop; MDI, multiple daily injections; no., number; SAP, sensor augmented pump

**Table 2:**
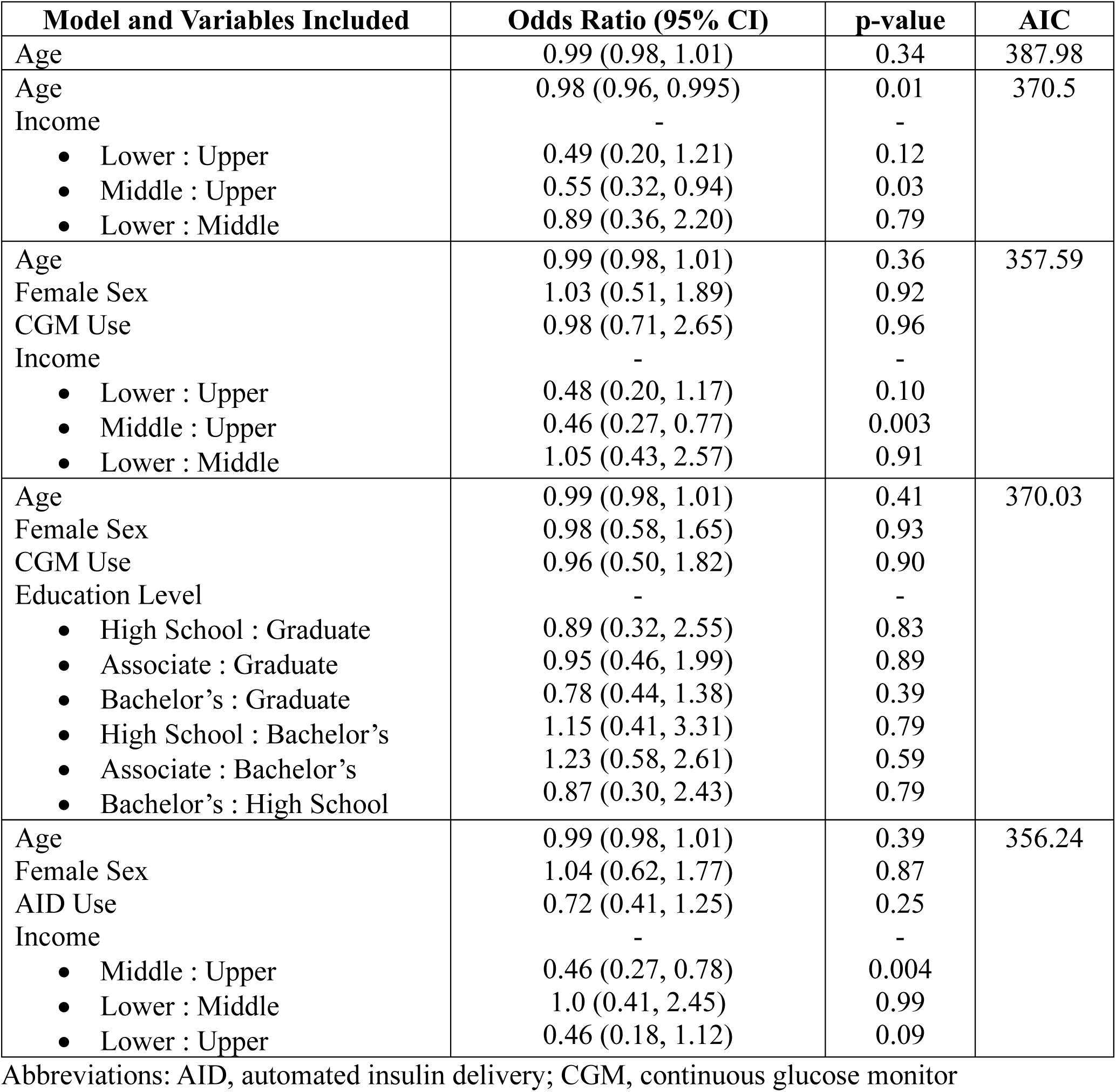
Multivariate logistic regression models for predicting target physical activity.

| **Model and Variables Included** | **Odds Ratio (95% CI)** | **p-value** | **AIC** |
| --- | --- | --- | --- |
| Age | 0.99 (0.98, 1.01) | 0.34 | 387.98 |
| Age | 0.98 (0.96, 0.995) | 0.01 | 370.5 |
| Income | - | - |  |
| • Lower : Upper | 0.49 (0.20, 1.21) | 0.12 |  |
| • Middle : Upper | 0.55 (0.32, 0.94) | 0.03 |  |
| • Lower : Middle | 0.89 (0.36, 2.20) | 0.79 |  |
| Age | 0.99 (0.98, 1.01) | 0.36 | 357.59 |
| Female Sex | 1.03 (0.51, 1.89) | 0.92 |  |
| CGM Use | 0.98 (0.71, 2.65) | 0.96 |  |
| Income | - | - |  |
| • Lower : Upper | 0.48 (0.20, 1.17) | 0.10 |  |
| • Middle : Upper | 0.46 (0.27, 0.77) | 0.003 |  |
| • Lower : Middle | 1.05 (0.43, 2.57) | 0.91 |  |
| Age | 0.99 (0.98, 1.01) | 0.41 | 370.03 |
| Female Sex | 0.98 (0.58, 1.65) | 0.93 |  |
| CGM Use | 0.96 (0.50, 1.82) | 0.90 |  |
| Education Level | - | - |  |
| • High School : Graduate | 0.89 (0.32, 2.55) | 0.83 |  |
| • Associate : Graduate | 0.95 (0.46, 1.99) | 0.89 |  |
| • Bachelor's : Graduate | 0.78 (0.44, 1.38) | 0.39 |  |
| • High School : Bachelor's | 1.15 (0.41, 3.31) | 0.79 |  |
| • Associate : Bachelor's | 1.23 (0.58, 2.61) | 0.59 |  |
| • Bachelor's : High School | 0.87 (0.30, 2.43) | 0.79 |  |
| Age | 0.99 (0.98, 1.01) | 0.39 | 356.24 |
| Female Sex | 1.04 (0.62, 1.77) | 0.87 |  |
| AID Use | 0.72 (0.41, 1.25) | 0.25 |  |
| Income | - | - |  |
| • Middle : Upper | 0.46 (0.27, 0.78) | 0.004 |  |
| • Lower : Middle | 1.0 (0.41, 2.45) | 0.99 |  |
| • Lower : Upper | 0.46 (0.18, 1.12) | 0.09 |  |

Abbreviations: AID, automated insulin delivery; CGM, continuous glucose monitor

### Facilitators and Barriers to Physical Activity

Reported barriers differed by those meeting and not meeting PA targets (Figure 1). In those meeting PA recommendation, the most common barrier, selected as likely to extremely likely to prevent activity, by 40% of participants was risk of hypoglycemia. The other barriers were uncommon and reported by less than 25% of those participants. For those not meeting PA target, risk of hypoglycemia was again the most common barrier (63%). Other common barriers in those not meeting PA target included weather conditions (52%), too tried to start (45%), other health issues not related to diabetes (41%), fear of losing T1D control (39%), cost of exercise (38%), tiring quickly (38%), and location of gym or exercise facility (26%). Each of these barriers differed between those meeting and not meeting PA targets (p<0.001).

**Figure 1.**
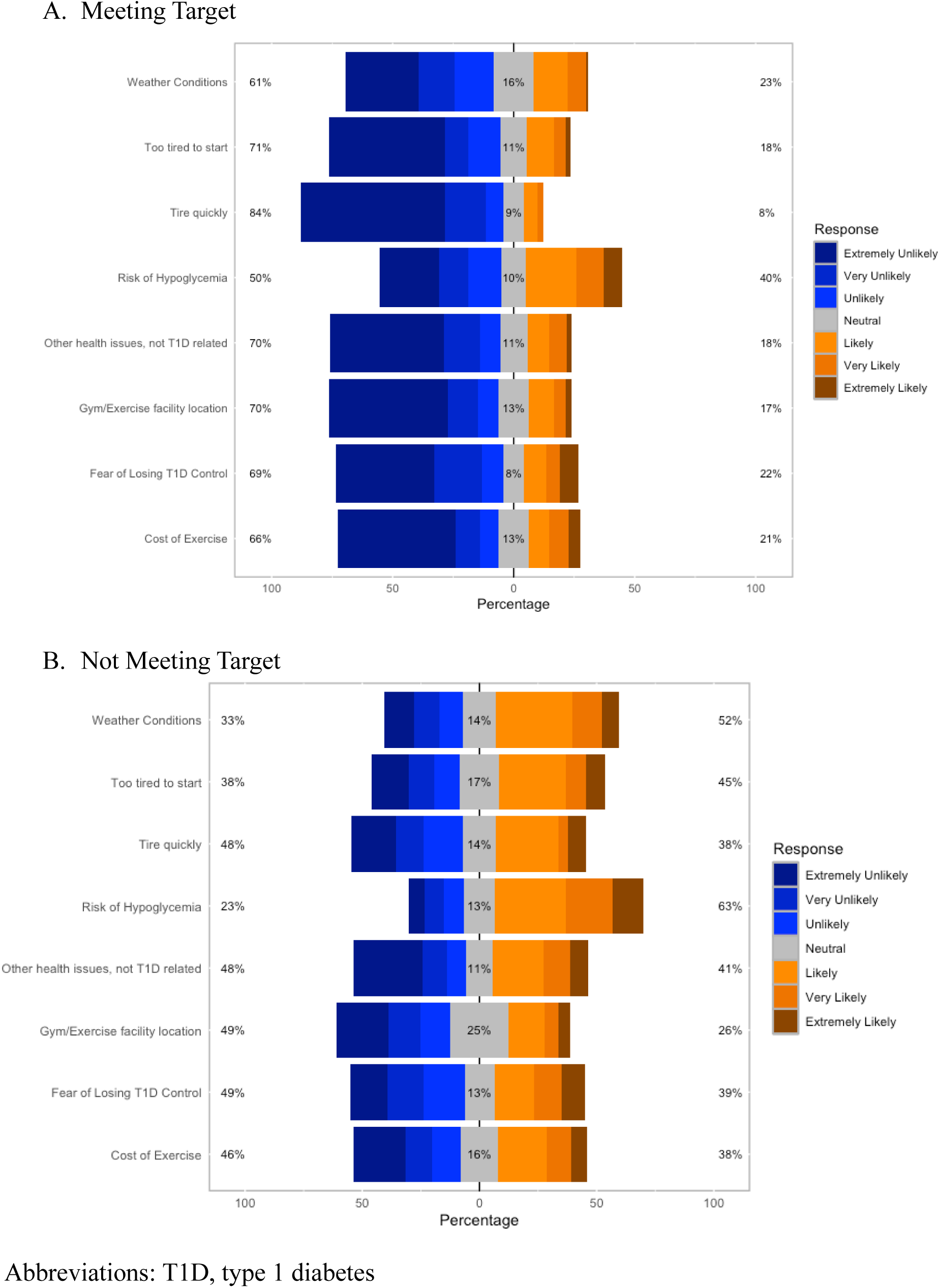
Distribution of Likert Scale Responses for Barriers to Physical Activity

Several facilitators to PA were chosen by over a third of participants: insurance reimbursing exercise program/gym (total sample: 50.9%, below target: 47.7%, meeting target: 53.6%), attending an exercise group and fitness advisor (total sample: 39.9%, below target: 38.5%, meeting target: 41.1%), and one-on-one advice from a health and fitness advisor (total sample: 36.4%, below target: 35.4%, meeting target: 39.1%). There were no differences within the selected facilitators between those meeting and not meeting PA targets (all; p>0.05).

Qualitative responses from participants with key themes are listed in Tables 3 and 4 (to questions: “What is the one thing that would most help you increase your exercise amount or intensity?” and “Is there anything that prevents you from exercising more?”) and online supplemental material Tables 1-3 (to questions: “What, if any, impact does diabetes have on your exercise habits?”, “What would help or encourage you to exercise more?”, and “Are there any other supports that you would be interested in or find helpful that are not listed above?”). The most reported barrier in qualitative responses related to lack of time. Approximately one quarter of participants reported that diabetes further complicated time barriers due to inflexibility related to insulin on board, a perceived need to exercise in the morning to prevent nocturnal hypoglycemia, and glucose levels below a target needed to initiate exercise. Concerns related to hypoglycemia were the second most common thematic code preventing exercise. This manifested as skipping activity due to glucoses below target or reluctance to consume calories to reach an optimal glycemic threshold prior to exercise which was perceived as counterproductive from a weight-management perspective. Participants also described the failure of current treatment modalities to adequately control blood glucoses during and after exercise. Universal barriers such as physical health conditions outside of diabetes, lack of motivation, and fatigue were also frequently described.

**Table 3:** Summary of responses to, “What is the one thing that would most help you increase your exercise amount or intensity?”

| **Key themes** | **Illustrative Response/Summary of Responses** |
| --- | --- |
| Consistent routine/time availability | *“Being able to exercise any time of day without having to eat more food in order to safely do it without risking low blood sugars”* |
| Peer accountability/support | These responses expressed a desire for peer accountability groups or some form of accountability and support in general. |
| Better control of blood sugar | *“Better ability to control for lows, especially around unplanned exercise (needing to run around at work, or deciding to go for a walk on a lunch break)”* |
| Better overall physical health/better energy levels | *“Not having chronic pain and having energy. I know that more exercise would be a good thing. But it has never been MY thing, I wish that it had been a life priority from when I was younger.”* |
| Motivation | *“Not sure how I can get this externally, but a positive attitude is essential for exercising. If I'm depressed about something or distressed because my blood sugar numbers haven't been what they should be through no fault of my own (gastroparesis, gluten contamination, stress, hormones, etc) that's when I might skip exercising.”* |
| Financial Assistance | *“I'd increase my variety of exercise if insurance paid for some of my classes/memberships”* |
| Professional support: medical or fitness | *“A trainer to provide advice about a plan to increase my intensity.”* |
| Fitness catered specifically to individuals with diabetes | *“Having a person with me who understands type 1 diabetes and what it can do to me while I exercise. Who also knows how to help me if I can't speak.”* |

**Table 4:**
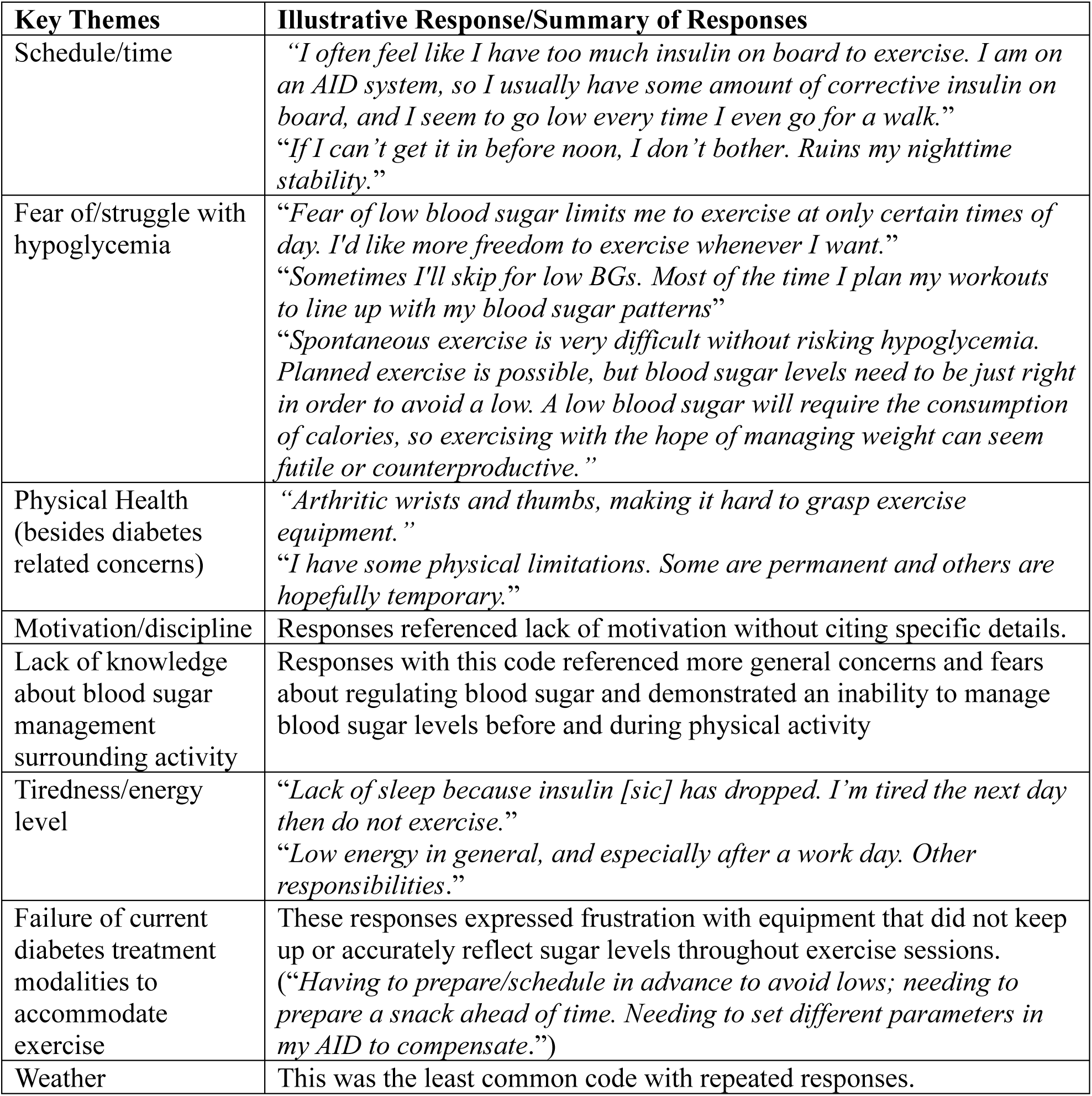
Summary of responses to: We know many people do not exercise as much as they would like to or feel they should. Is there anything that prevents you from exercising more?

Regarding qualitative responses to PA facilitators, the most common theme was greater flexibility. Participants desired the ability to have unplanned exercise, suggesting the following solutions in free responses: more advanced predictive AID algorithms, more facile insulins, or additional opportunities to exercise in the morning, which many respondents felt was the optimal time to control glucose and avoid nocturnal hypoglycemia. In addition, a preference for peer support or accountability was commonly reported. Many further indicated a desire for fitness tailored to people with type 1 diabetes specifically involving peers or professionals knowledgeable about type 1 diabetes glycemic management. Financial assistance and/or insurance reimbursement for full or partial gym membership were also described as facilitators. Finally, some participants expressed desire for education about glycemic management surrounding exercise and assistance from health professionals to optimize pump settings during PA.

### Diabetes Quality of Life (DQOL)

There were no differences in DQOL scores between those that did (12.0 ± 3.96) and did not (11.9±4.64) meet PA targets (p=0.43).

### Multivariable logistic regression

Multivariable logistic regression was used to determine predictor variables independently associated with target PA. The results of each model are listed in Table 2. The model that was the best fit according to AIC adjusted for age, sex, AID use, and income level (AIC: 356.24). Middle versus upper income level was associated with 54% lower odds of achieving target physical activity (adjusted odds ratio [aOR]: 0.46, 95% confidence interval [CI]: 0.27—0.78, 0.004). A similar effect size was observed comparing those at the lowest to highest income level (aOR: 0.46, 95% CI: 0.18—1.12, p=0.09) although that was not statistically significant. Education level was not significantly associated with target PA adjusting for age, sex, and CGM use.

## Discussion

This research has several key findings which contribute to our understanding of barriers and facilitators to PA for people with type 1 diabetes living predominately in the U.S. One of the most consistent themes across all domains (quantitative and qualitative data) related to cost of PA. The most selected PA facilitator for people with type 1 diabetes was insurance reimbursing an exercise program or exercise facility membership. In quantitative and qualitative responses, cost was reported as a barrier more frequently than access to an exercise facility, and some respondents conveyed that reimbursement of even a portion of gym membership costs could facilitate their activity. This was also in line with the multivariable logistic regression results in which the highest income group was more likely to reach target PA even after adjusting for age, sex, and use of either a CGM or AID device. Interestingly, sex, age, and use of diabetes technology or education level were not predictive of target PA. Although only the highest to middle income comparison was significant, the lowest to highest income comparison had a similar effect size but may have been underpowered in the lowest income group to detect a significant difference. The consistency of the results across several domains underscores the importance of future studies interrogating the cost-efficacy of insurance reimbursement of type 1 diabetes exercise programs or facility memberships. Although a cost effectiveness analysis has not yet been performed in type 1 diabetes, most exercise interventions are cost effective or cost saving for people with type 2 diabetes (23–25). It stands to reason that exercise programs would be similarly cost effective for people with type 1 diabetes.

Other facilitators included fitness tailored to individuals with type 1 diabetes and peer support, as one respondent explained, “Having a person with me who understands type 1 diabetes and what it can do to me while I exercise. Who also knows how to help me if I can’t speak.” Turning to the limited intervention evidence from the U.S. type 1 diabetes population, two adolescent group physical activity programs, conducted in person or with virtual exercise games, demonstrated safety and feasibility (26,27). These programs could serve as models for larger studies with wider age eligibility.

It is also worth noting that risk of hypoglycemia was the most selected barrier to PA in all participants, regardless of reaching or not reaching target PA levels. Nevertheless, participants not reaching target PA rated this as a more likely barrier. Qualitative responses detailed the many facets whereby the threat of hypoglycemia prevented exercise, and these responses could serve as intervention opportunities for healthcare providers. Some participants perceived that consuming glucose containing calories to reach acceptable glucose thresholds for activity counteracted physical activity benefits. Many participants perceived a need to perform PA early in the day to prevent nocturnal glucose instability. Some participants also indicated that optimization of insulin regimens for PA or education regarding glucose management surrounding exercise would support activity. These responses suggest that further education regarding health benefits of PA for people with type 1 diabetes and routine discussion of a plan for glucose management before, during, and after (including prevention of nocturnal hypoglycemia) might promote PA. A general dissatisfaction with the capacity of diabetes treatment modalities, including state-of-the-art AID systems, to support unplanned exercise was another consistent perception, demonstrating an unmet need for further technology and pharmaceutical development to support healthy behaviors.

This study builds on prior qualitative research examining barriers and facilitators to PA in type 1 diabetes, adding to the literature a larger population of a predominantly U.S-based cohort including individuals meeting PA recommendations. Most prior studies in type 1 diabetes similarly reported actual or fear of hypoglycemia as a major barrier to PA including initiation or reaching desired duration or intensity (12–14,17). Although cost *per se* is not a previously identified barrier, limited access to exercise facilities was previously reported (13). As demonstrated in the current study, education and additional peer or healthcare provider support were consistently observed facilitators in prior studies (12,13,15–17). While major themes have recurred between studies, it is also worth noting the variability of responses even within studies. This heterogeneity underlines the importance of healthcare providers using open-ended questions in discussions regarding PA barriers with patients living with type 1 diabetes as an opportunity to elicit personalized concerns and provide education where appropriate or troubleshoot modifiable barriers.

This study has several limitations. The participants recruited were predominately white, non-Hispanic and most had achieved advanced education levels. This limits broader study generalizability. The online-based survey method inherently restricted participants to those with computer and internet access. We also recruited participants from organizations and websites associated with type 1 diabetes, which may have led to an over-representation of participants with high type 1 diabetes knowledge and support engagement. This is corroborated by the fact that a vast majority of participants understood that PA is beneficial for people with type 1 diabetes and were established with an endocrinologist. For future studies, use of methods not requiring internet or computer and recruiting a more socioeconomically, racially, and ethnically diverse population through community engagement or in-person recruitment from medical centers could enhance result generalizability. Additionally, because the survey was anonymous, which had the advantage of increasing the likelihood of responding to sensitive questions while protecting confidentiality, we could not collect data on the number of participants who viewed the recruitment materials or verify that each participant provided only one survey response.

Finally, physical activity intensity and duration data were provided via self-report instead of through objective measurement, which could lead to inaccurate reporting (28). Despite these limitations, this study has several strengths including a large response from a previously under-studied population across the U.S., mixed methodologies with quantitative and qualitative components, and the ability to examine in depth demographic characteristics including income, education, and insurance status.

In conclusion, this survey-based study identified cost as a newly reported barrier to PA for people with type 1 diabetes living predominately in the U.S. and reimbursement of exercise programs or facility memberships as a facilitator. Many participants also desired technological/pharmacological advances to support flexible PA, education about glucose management surrounding PA, and PA tailored to people with type 1 diabetes through peer or professional support to facilitate engagement.

## Data Availability

All data produced in the present study are available upon reasonable request to the authors

## Acknowledgements

Participants were recruited from the T1D Exchange Registry – an online registry of individuals with type 1 diabetes and caregivers officially launched in 2019. We thank the participants of the T1D Exchange Registry and all people with type 1 diabetes who participated for their time and interest in this study.

