## Supplementary material for "Financial factors: a mixed methods survey-based study of barriers and facilitators to physical activity in type 1 diabetes": Online Supplemental Fig. 1; online supplemental material Tables 1-3

Online-Only Supplemental Material

**Supplemental Figure 1. FLOW Diagram**

306 Participants

No Physical Activity Response

(n=25)

281 Complete Response

**Supplemental Table 1: Summary of responses to, “What, if any, impact does diabetes have on your exercise habits?”**

| **Key themes** | **Illustrative Response/Summary of Responses** |
| --- | --- |
| Fear or consequences of hypoglycemia | *“Diabetes has always limited exercise. There is so much work to manage this disease to exercise that it is often not worth it. For example, I love Pilates but have trouble joining a class and then driving home without getting low. A tricky balancing act.*” |
| Timing/scheduling/lack of spontaneity | “*So much planning is necessary in order to do it safely. Did I reduce my IOB enough ahead of time? Do I have rescue carbs and Baqsimi with me? Am I hydrated enough?*” |
| No concerns/I have learned to adapt to these impacts | This was the third most popular code for this question. |
| Glucose-related concerns outside of hypoglycemia | “*I am having to constantly monitor blood glucose levels and take unplanned breaks to allow lows to be treated. I also need to plan ahead to set exercise mode on my pump a few hours ahead to get optimal control over blood glucose levels. I feel some of the benefits of the exercise are lost due to having to take glucose tabs to treat lows in order to complete a workout.*” |
| Intensity/types of exercise | “*It limits the intensity, duration and flexibility of when to exercise.*” |
| Food/meal planning/eating beforehand | “*Have to think about when I have no insulin on board, have to eat a lot beforehand, have to avoid HIIT workouts.*” |
| Incentive/Diabetes has encouraged me to exercise more | “*It motivates me to exercise more. Prior to being diagnosed at the age of 42, I went through spurts of exercise. Now, I am motivated to do more because I can see the exact consequences of my actions. If I stop exercising, my diabetes quickly gets high and stays there*.” |

**Supplemental Table 2: Summary of responses to: “What would help or encourage you to exercise more?”**

| **Key Themes** | **Illustrative Response/Summary of Responses** |
| --- | --- |
| Nothing/I don’t need to exercise more | The most common code. Responses largely echoing themes of being satisfied with their current exercise routine |
| Help with scheduling/time/ flexibility | “*A more routine schedule.*”  “*More flexibility in early morning group exercise classes,*” as many had previously stated that morning workouts were the best way to maintain blood sugar levels. |
| Better diabetes treatments (to prevent hypoglycemia or glucose instability during exercise) | “*More reliable blood sugar numbers during the workout. For example, sometimes my BG goes up while working out, then immediately falls afterwards.”*  “*Knowing my blood sugars would not spike afterwards or dive during. Being in that balance is scary for me and my family members*.”  “*I need to retest all my pump settings. Also I think it would be helpful to talk to a PWD [sic] who is knowledgeable about exercise physiology.”* |
| Peer accountability and group/technology support | “*An accountability partner or something that would help remind me to move. Also, something like an app that can give me suggestions for movement on the go*.” |
| Personalized workouts or groups geared toward people with diabetes | “*A diabetes exercise support group that logs exercise*.”  “*Having people to do it with me, a trainer to teach me how to lift weights*”. |
| Better physical health | “*Well, if I was capable of it, I'd exercise a lot more. I enjoy a variety of cardio activities from running, biking, skiing, and such*.” |
| Education/Better understanding of sugar management | *“Better pump settings and more education.”*  *“To be taught more on how to manage my diabetes with high intensity exercise, instead of just being given the advice to eat a sugary snack in the event of a hypo.”* |

**Supplemental Table 3: Summary of responses to: “Are there any other supports that you would be interested in or find helpful that are not listed above? Please describe.”**

| **Key Themes** | **Illustrative Response/Summary of Responses** |
| --- | --- |
| None | This was the most popular code for this question. |
| Peer support (Accountability partners, organized group support/classes) | This was, by far, the next most popular code for this question by nearly 3 to 1. Some expressed a desire for general peer supports but many expressed a desire for support tailored to people with type 1 diabetes: “*I would love "diabetic specific" info and guidance. Everything I know about working out as a diabetic right now is stuff I've read online from who-knows-who (e.g. people may or may not be professionals)”* |
| Financial assistance | *“Perhaps if a free gym membership is not possible then potentially a subsidized membership given the importance of exercise to managing Type I diabetes and maintaining good health.”* |
| Better diabetes treatments | “*Better, faster reacting insulins.*”  “*Better integration of exercise data with pump and cgm data, including AI analysis*.” |
| Medical support | These responses specifically mentioned the desire to work with a physical therapist or general practitioner to support regular exercise. |
| Facility accessibility | These responses described lack of physical facilities (gyms, etc.) in close proximity. |
