## Supplementary material for "Financial factors: a mixed methods survey-based study of barriers and facilitators to physical activity in type 1 diabetes": Online Supplemental Material

****[SECTION HEADER] Thank you for taking this survey. This study wants to learn about the health habits and views of people with type 1 diabetes. Your participation is greatly appreciated.

Have you been told by a doctor or other healthcare professional that you have type 1 diabetes?

[ ] Yes

[ ] No

Are you 18 years or older?

[ ] Yes

[ ] No

[If no to above questions, the following message will appear: We appreciate your interest in the survey, but we are only surveying people with type 1 diabetes who are 18 or older.]

Age (years) __________________________________

How many years ago were you diagnosed with type 1 diabetes? If less than 1 year, enter 0. __________________________________ years

Have you seen an endocrinologist, a doctor or other healthcare provider who specializes in diabetes management, in the past 3 years?

[ ] yes

[ ] no

[ ] Not sure/prefer not to say

What do you use to manage your diabetes (check all that apply)?

[ ] Long and short acting insulin shots

[ ] Continuous glucose monitor or CGM (For example Libre, Dexcom, Guardian, Eversense sensors)

[ ] Insulin pump without sensor input (I do not have a CGM or it does not connect to my pump to adjust insulin)

[ ] Insulin pump with suspend before low (my pump reduces or stops insulin to prevent low blood sugars)

[ ] Hybrid closed loop insulin pump system (my pump increases and decreases insulin based on blood sugars)

[ ] Other (please specify) _____________________________________

***[SECTION HEADER] This next section asks about exercise or physical activity. Exercise or physical activity can vary by level of intensity. For each activity level below, please select the approximate number of minutes in a typical week that you spend in this activity level.

About how many minutes per week do you spend in light physical activity (you are breathing normally and comfortably)? For example, walking slowly around a store or walking the dog:

[ ] None

[ ] 5-29 minutes (under half an hour)

[ ] 30 to 59 minutes (half an hour to just under 1 hour)

[ ] 60 to 89 minutes (1 hour to just under 1 hour and a half)

[ ] 90 to 119 minutes (1 hour and a half to just under 2 hours)

[ ] 120 to 149 minutes (2 hours to just under 2 and a half hours)

[ ] 150 to 179 minutes (2 and a half hours to just under 3 hours)

[ ] 180 minutes or more (3 hours or more)

About how many minutes per week do you spend in moderate intensity physical activity (your breathing quickens but you are not out of breath, you develop a light sweat after 10 minutes, you can carry on a conversation). For example, walking briskly at the pace that you are late for a meeting:

[ ] None

[ ] 5-29 minutes (under half an hour)

[ ] 30 to 59 minutes (half an hour to just under 1 hour)

[ ] 60 to 89 minutes (1 hour to just under 1 hour and a half)

[ ] 90 to 119 minutes (1 hour and a half to just under 2 hours)

[ ] 120 to 149 minutes (2 hours to just under 2 and a half hours)

[ ] 150 to 179 minutes (2 and a half hours to just under 3 hours)

[ ] 180 minutes or more (3 hours or more)

****[SECTION HEADER] The next questions ask about different types of activity. Please note, the times listed are different from the prior questions.

About how many minutes per week do you spend in high intensity physical activity (your breathing is deep and rapid, you can't say more than a few words without pausing for breath, you develop a sweat after only a few minutes)? For example, running:

[ ] None

[ ] 1 to 8 minutes

[ ] 9 to 29 minutes

[ ] 30 to 49 minutes

[ ] 50 to 69 minutes

[ ] 70 to 89 minutes

[ ] 90 minutes or more

About how many minutes per week do you spend strength training? For example, using weights, doing squats or push-ups:

[ ] None

[ ] 1 to 8 minutes

[ ] 9 to 29 minutes

[ ] 30 to 49 minutes

[ ] 50 to 69 minutes

[ ] 70 to 89 minutes

[ ] 90 minutes or more

Do you participate in any other activities not listed? (please specify activity and time spent): __________________________________ (minutes per week)

Where do you typically go to engage in recreational physical activity, like exercise or sports, during your free time?

- None, not currently active
- Local park
- Fitness center/gym
- Community recreational center
- Hiking trails
- Sports leagues/fields
- Swimming pool
- Home gym
- Local walking paths
- Other (*please specify*): _________________________________

Do you currently work?

[ ] yes

[] no

[If yes]: How active are you during your usual work day?

- I am sedentary most of the day (for example, sitting at a desk, chair, etc)
- I move for about half the day
- I am on my feet but standing still (for example, working from a standing desk)
- I am constantly walking, moving, or lifting
- Other, please specify: _________________________

***[SECTION HEADER] This next set of questions asks about diabetes and how it impacts your life.

Please rate each statement below from very satisfied (1) to very dissatisfied (5).

How satisfied are you with your current diabetes treatment?

1) Very satisfied
2) Moderately satisfied
3) Neither satisfied nor dissatisfied
4) Moderately dissatisfied
5) Very dissatisfied

How satisfied are you with the amount of time it takes to manage your diabetes?

1) Very satisfied
2) Moderately satisfied
3) Neither satisfied nor dissatisfied
4) Moderately dissatisfied
5) Very dissatisfied

How satisfied are you with the time you spend exercising?

1) Very satisfied
2) Moderately satisfied
3) Neither satisfied nor dissatisfied
4) Moderately dissatisfied
5) Very dissatisfied

Please rate the following questions from never (1) to all the time (5).

How often do you worry about whether you will miss work/school?

1) Never
2) Very seldom
3) Sometimes
4) Often
5) All the time

How often do you have a bad night’s sleep because of diabetes?

1) Never
2) Very seldom
3) Sometimes
4) Often
5) All the time

How often do you feel diabetes limits your career?

1) Never
2) Very seldom
3) Sometimes
4) Often
5) All the time

How often do you worry about passing out?

1) Never
2) Very seldom
3) Sometimes
4) Often
5) All the time

***[SECTION HEADER] The next set of questions ask about barriers to exercise for people with type 1 diabetes:

We know many people do not exercise as much as they would like to or feel they should. Is there anything that prevents you from exercising more? ______________________________________________________________________________________________________________________________________________________________________________________________________________________________________________________________________________________________________

What would help or encourage you to exercise more? __________________________________________

What, if any, impact does diabetes have on your exercise habits? __________________________________________

Do you think exercise is beneficial for people with type 1 diabetes? (select one)

- Yes
- No

[If no] Please explain why exercise is **not** beneficial to people with type 1 diabetes and how you learned this or came to this understanding. ______________________________________________________________________________________________________________________________________________________________________________________________________________________________________________________________________________________________________

***[SECTION HEADER] The list below includes different reasons given by some people with type 1 diabetes for **not** engaging in physical activity. For each one, please tell us the likelihood, from extremely unlikely (1) to extremely likely (7), that it would **stop you** from engaging in regular physical activity during the next 3 weeks:

1. The loss of control over your diabetes

2. The risk of low blood sugars

3. The fear of being tired

4. The fear of hurting yourself

5. The fear of suffering a heart attack

6. A low fitness level

7. The fact that you have diabetes

8. The risk of high blood sugars

9. Health issues outside of your diabetes

10. Weather conditions

11. The location of a gym or other exercise place

12. Your work schedule

13. Family obligations

14. Cost of a gym or other place to exercise

15. Concern that exercise is not healthy for people with diabetes

16. You are too worn-out to start exercise

17. You feel tired very quickly during exercise

18. You don’t feel comfortable adjusting insulin or diet to manage blood sugars around activity times

19. Other factors that would keep you from exercise (please specify):

***[SECTION HEADER] Listed below are types of supports that may help people start exercising or increase the amount they exercise. What type(s) of support would **you** be interested in or have found helpful?

Please select all that apply.

- one-on-one advice from a health and fitness advisor
- attending an exercise group (for example, fitness class or meeting with a group of individuals to walk/run/bike/swim together)
- motivational support through a professional who keeps in touch to see how you are doing with your exercise program
- counseling by your endocrinologist or other doctor about managing blood sugars around the time of exercise
- using telehealth (meeting with a fitness coach remotely by phone or computer) during your exercise sessions where you can participate from home or at a location convenient to you
- insurance paying for an exercise program or having free access to a gym or exercise program
- other (please specify below)
- none of these

Are there any other supports that you would be interested in or find helpful that are not listed above? Please describe: ________________________________

What is the one thing that would **most** help you increase your exercise amount or intensity?___________________

****[SECTION HEADER] Thank you for your responses. This is the last set of questions, and they give us general information about your background. As a reminder, all information is kept private and confidential.

How do you describe yourself?

- Man
- Woman
- Non-binary
- Prefer to provide my own description: ________________________________
- Prefer not to answer

How do you describe yourself? *Please select all that apply*:

- American Indian or Alaskan Native
- Asian or Asian American
- Black or African American
- Hispanic/Latinx
- Middle Eastern/North African
- Native Hawaiian/Pacific Islander
- White or Caucasian
- Prefer to provide own description: ______________________________
- Prefer not to answer

Which state do you live in? (Drop down list of states)

How would you describe the area where you live?

- Urban (city)
- Suburban (just outside of a city)
- Rural (outside of a major city, in the countryside)
- Other (*please specify*):______________________

What is your primary source of healthcare coverage? *That is, the insurance you use most often*.

- Private health insurance through your or someone else’s employer or union
- Medicare
- Medicaid
- Health insurance that you bought from a state or federal Insurance Marketplace
- Military/VA/TriCare
- Other (please specify): ______________________________
- I don’t currently have any healthcare coverage
- I’m not sure
- Prefer not to answer

What is the highest grade or level of school you completed?

- Up through grade 8 or less
- Some high school
- High school graduate/GED
- Associate’s degree, CTE certificate, or some college
- Bachelor’s degree
- Some graduate work
- Master’s, Professional, or Doctoral degree
- Other (please specify): _____________________
- Prefer not to answer

This past year, was your annual household income from all sources:

- Less than $10,000
- $10,000 - $14,999
- $15,000 - $19,999
- $20,000 - $34,999
- $35,000 - $49,999
- $50,000 - $74,999
- $75,000 - $99,999
- $100,000 - $149,999
- More than $150,000
- Don’t know
- Prefer not to answer

[If survey submitted] Thank you for completing the survey! Your participation is appreciated.
